# Serological assessment of SARS-CoV-2 exposure in northern Sweden by the use of at-home sampling to meet geographical challenges in rural regions

**DOI:** 10.1101/2020.06.02.20120477

**Authors:** Julia Wigren Byström, Linnea Vikström, Ebba Rosendal, Remigius Gröning, Yong-Dae Gwon, Emma Nilsson, Atin Sharma, Akbar Espaillat, Leo Hanke, Gerald McInerney, Andrea Puhar, Felipe Cava, Gunilla B Karlsson Hedestam, Therese Thunberg, Tor Monsen, Fredrik Elgh, Magnus Evander, Anders Johansson, Anna K Överby, Clas Ahlm, Johan Normark, Mattias NE Forsell

**Affiliations:** Department of Clinical Microbiology, Umeå University, Swedem; Department of Xerum AB, Sweden, Umeå University, Sweden; Department of Molecular Biology, Umeå University, Sweden; Department of Laboratory for Molecular Infection Medicine Sweden, Umeå University, Sweden; Department of Microbiology, Tumor and Cell Biology, Karolinska Institutet, Sweden

## Abstract

The current SARS-CoV-2 pandemic has highlighted a need for easy and safe blood sampling in combination with accurate serological methodology. Venipuncture is usually performed by trained staff at health care centers. Long travel distances may introduce a bias of testing towards relatively large communities with close access to health care centers. Rural regions may thus be overlooked. Here, we demonstrate a sensitive method to measure antibodies to the S-protein of SARS-CoV-2. We adapted and optimized this assay for clinical use together with capillary blood sampling to meet the geographical challenges of serosurveillance. Finally, we tested remote at-home capillary blood sampling together with centralized assessment of S-specific IgG in a rural region of northern Scandinavia that encompasses 55,185 sq kilometers. We conclude that serological assessment from capillary blood sampling gives comparable results as analysis of venous blood. Importantly, at-home sampling enabled citizens living in remote rural areas access to centralized and sensitive laboratory antibody tests.

## INTRODUCTION

The highly contagious virus SARS-CoV-2 has caused a world-wide pandemic with severe consequences for individuals and societies across the globe. Real-time monitoring of how the virus spreads is achieved by extensive testing and analysis of virus RNA in respiratory samples. Serological studies have also been of importance, as previous exposure reduces the risk of developing severe or fatal COVID-19.(1, 2) It is clear that real-time epidemiological sero-surveillance is critical to support governmental decision making. On an individual level, seroconversion to virus-specific IgG reflects the development of virus-specific B and T cell memory.(3) Accordingly, information of seroconversion can be used to estimate likelihood of severe disease in a population, as well as to devise and monitor strategies to reduce further cases of COVID-19. This includes the allocation and prioritizing of healthcare resources and vaccine rollout.(4-6)

It is widely accepted that most patients with clinical COVID-19 have developed robust isotype-switched antibody-responses to SARS-CoV-2 between 1-2 weeks of disease onset(7, 8), and these are readily detected by many currently available point-of-care or laboratory-based serological assays.(9) However, a large majority of all SARS-CoV-2 infections are asymptomatic or manifest with mild disease(10, 11), and not all individuals develop strong and uniform antibody responses to all proteins of SARS-CoV-2. The Spike protein (S) of SARS-CoV-2 mediates attachment and entry of the virus into target cells(12, 13). Antibodies that can block these events are neutralizing, and there is a good correlation between S-binding antibodies and capability of immune serum to neutralize the original Wuhan-1 isolate of SARS-CoV-2 and variants of concern preceding the current Omicron strain(14). Vaccine-induced immune responses to the S-protein also forms the basis of currently used Covid-19 vaccines. As such, methodologies that measure antibodies towards S can be used both for serosurveillance of infections and for assessment of vaccine-induced responses.

Here, we describe the development and evaluation of a sensitive assay that was optimized for detection of early or low-level anti-SARS-CoV-2 IgG responses and that was implemented in clinical practice at the University Hospital of Northern Sweden. We also demonstrate the feasibility of remote sampling in combination with highly sensitive diagnostics in a sparsely populated geographic region of northern Sweden. Collectively, the findings reported here provide a blueprint for population-based immunosurveillance of SARS-CoV-2 exposure or vaccine coverage that is not limited by geographical constraints.

## METHODS

### Human samples and ethical permission

All serum samples within this study were collected for clinical purposes within the realm of providing healthcare to patients in Region Västerbotten, Sweden and stored at the Department of Laboratory Medicine, Umeå University Hospital. At-home sampling kits were offered to all inhabitants of Västerbotten that were over the age of 16. After return of kits to Norrland’s University Hospital, analysis was done with regards to successful sampling and S-directed reactivity by the Regional Laboratory. Test results were returned to the individual under confidentiality in accordance with clinical practice. Anonymized data for evaluation of the at-home sampling or venous blood sampling strategies were provided by Region Västerbotten.

For comparison, we investigated 30 samples from 25 hospitalized COVID-19 patients with moderate to severe disease and 144 samples from before the emergence of SARS-CoV-2. In addition, samples that had been collected for the purpose of infection prevention at a care home for the elderly were used in this study. Ethical permission for the study was obtained from the Swedish Ethical review authority (No: 2020-01557) and all research was carried out according to The Code of Ethics of the World Medical Association (Declaration of Helsinki).

### Protein production

Plasmid encoding the 2019-nCoV S protein was kindly provided by Jason McLellan and the details of these constructs has been previously described(15). Protein was produced using the Freestyle™ MAX 293 Expression System (Thermo Fisher Scientific). Briefly, plasmids encoding the SARS-CoV-2 S protein (Wuhan strain) was transfected into the cells using Freestyle™ MAX reagent in a 1×10^6^ cells/ml culture and grown for 4-5 days in 8% CO_2_ and 120 rpm. The supernatant was then cleared of cells and debris by centrifugation and by passage through a 0.2 uM filter. The supernatant was then flowed over column packed with His-pure Ni-NTA resin (Thermo Fisher Scientific) at a rate of approximately 0.5ml/minute. Subsequently the column was washed with 10 column volumes of 20mM Imidazole / PBS pH 7.4 and then eluted with 250mM Imidazole / PBS pH 7.4. The resulting elute was concentrated and buffer exchanged to PBS pH 7.4 by Amicon spin columns with a cut off <100kDa (Sigma Aldrich). In an alternative set-up for purification of S protein for competition assays, the S protein was first captured via glycans in lentil-lectin cromatography (GE Healthcare), washed with PBS and then eluted with 0.5M Methyl-α-D-mannopyranoside prior to loading of the elute on a Ni-NTA packed column. Protein purity was determined by SDS-page, native gel electrophoresis and concentration was determined by a BCA Protein Assay kit (Thermo Fisher Scientific).

### SARS-CoV-2 S ELISAs

In this study, we used two SARS-CoV-2 Spike ELISA protocols: For both assays, clear flat-bottom Immuno Maxisorp 96-well plates (Thermo Scientific) were coated with 200 ng/well of purified SARS-CoV-2 S protein and incubated at +4°C overnight. The following day the wells were washed once with PBS-0.05% Tween (PBS-T) and incubated for 1h at RT with blocking buffer (1% non-fat dry-milk in PBS-T). In the S ELISA protocol, duplicates of heat-inactivated human serum samples were diluted 1/100 in blocking buffer and added to the ELISA plate. For each plate, control serum samples from a highly anti-S IgG positive individual, and blank wells with blocking buffer alone was also included. Subsequently, the plate was incubated for 1h at RT and then washed four times with PBS-T. Goat-anti human IgG alkaline phosphate (AP)-conjugated antibody (Thermo Scientific) was diluted 1/6000 in blocking buffer and 100 µl was added to each well and incubated for 1 h at +37°C. The wells were then washed four times with PBS-T, followed by addition of 100µl/well of AP colorimetric substrate containing 1 mg/ml Phosphatase Substrate (Sigma-Aldrich) dissolved in diethanolamine buffer. The plate was incubated at +37°C for 30 min, and the reaction was stopped with 50 µl 3 M NaOH. Detection of anti-S IgG was then analyzed at 405nm with Tecan Sunrise reader. The final OD was calculated as OD_405nm_(sample) – OD_405nm_ (blank well). In some instances, we used a HRP-conjugated anti-human IgG as secondary antibody (Thermo Scientific) at a dilution of 1/5000 dilution in blocking buffer. Colorimetric change of 1-Step Ultra TMB-ELISA (Thermo Scientific) was measured at 450nm after 2M H2SO4 had been added to stop the reaction.

### Spike competition assay

A spike protein competition assay was performed to verify inconsistent positive SARS-CoV-2 antibody results that differed between the S ELISA and the Architecht or Liaison instruments. Serum was diluted 1/10 in PBS and purified S protein (100 µg/ml) or mock (PBS) was added and incubated for 1h at RT. The serum/S protein mixture was diluted ten-fold with blocking buffer to obtain a final serum dilution of 1/100 and 100 µl per well was added in duplicates to an S ELISA plate.

### qDBS sample preparation

After successful sampling, the sample card (qDBS, Capitainer AB) comprised 2 paper discs, each with 10µl of whole blood. The discs were extracted from the qDBS sample card by using a semi-automated puncher (qDBS Card Puncher, Capitainer AB) and one extracted disc was placed each in a single well of a 96-well plate. Blood components were then eluted from the disc by adding 100 µl PBS-T containing protease inhibitor cocktail (#4693116001, Roche) and subsequent incubation in a shaker (170rpm) for 1h in RT. Twenty µl blood eluate were then mixed with 80 µl blocking buffer and the eluate was analyzed in duplicates for SARS-CoV-2 S-directed IgG at a final dilution of 1/50.

### Stability test for seasonal changes

Anonymized positive- and negative control human plasma samples with known S-directed IgG levels were used to spike EDTA blood samples from a healthy donor that was negative for S-directed IgG. Spiked blood with negative (n=30), low (n=60), moderate (n=30) or high (n=30) S-directed IgG was then applied to qDBS sampling cards. The qDBS sampling cards were then put in paper envelopes and incubated at RT or in a climatic chamber (MHK-800 YK, Euroterm) that had been programmed with two different temperature and humidity scenarios, as specified by FDAs Guidance Document on Home Specimen Collection Serology Template for Fingerstick Dried Blood Spot (DBS) from the Policy for Coronavirus Disease-2019 Tests During the Public Health Emergency (https://www.fda.gov/regulatory-information/search-fda-guidance-documents/policy-coronavirus-disease-2019-tests-during-public-health-emergency-revised). These two different scenarios were used to reflect shipping of sample cards under winter or summer conditions.

### Virus and plaque neutralization assay

Vero E6 cells were cultured in Dulbecco’s modified Eagle’s medium (DMEM, D5648 Sigma) supplemented with 5 % FBS (HyClone), 10 units/mL penicillin and 10 µg/ml streptomycin (PeSt, HyClone). The patient isolate SARS-CoV-2/01/human/2020/SWE accession no/GeneBank no MT093571.1, was provided by the Public Health Agency of Sweden. The viral stock was grown in VeroE6 cells for 48 h and titrated by plaque assay. Plaque assay: VeroE6 cells (4×10^5^/well) were seeded in 12 well plates (VWR) 12-24 h prior to infection, and tenfold serial dilution of virus was added to the cells. We removed the virus inoculum after 1 h and added 2 ml semisolid overlay containing DMEM + 2 % FBS + PeSt + 1.2 % Avicel RC/CL and cells were incubated at 37 ºC in 5 % CO_2_. After 65 h the semisolid overlay was removed and cells were fixed with 4 % formaldehyde for 30 min. Cells were washed with PBS and stained with 0.5 % crystal violet in 20 % MeOH for 5 minutes. Plates were washed with water and the plaques counted. Plaque neutralization assay: Serum samples were heat inactivated at 56 ºC for 30 min prior to analysis. Ten-fold dilutions of sera were performed in virus stock (250 PFU/ml in DMEM + PeSt). The virus/sera mix was incubated at 37 ºC in 5 % CO_2_ for 1h, then 400 µl virus/sera inoculum was added to VeroE6 cells and plaque assay preformed as described above.

### Fluorescent inhibition assay

Vero E6 cells (10^4^/well) were seeded 12-24 h in 96 well plates (Greiner CELLSTAR^®^) prior to infection. Heat inactivated serum samples were diluted 1:10 in virus solution (10^4^ PFU/ml in DMEM + PeSt) and then further five-fold serial diluted in same virus preparation. The virus/sera mix was incubated at 37 ºC in 5 % CO_2_ for 30 min, then 50 µl virus/sera inoculum was added to the cells and incubated for another 2 h at 37 ºC in 5 % CO_2_. The inoculum was removed and 100 µl media containing DMEM + 2% FBS + PeSt was added and the cells were incubated at 37 ºC in 5 % CO_2_. 8 hours post infection the cells were prefixed by removing 50 µl of media and adding 50 µl of 4% formaldehyde for 10 min at RT followed by fixation for 30 minutes in 4 % formaldehyde. Plates were washed with PBS, permeabilized with 0.5 % Triton X-100 in PBS and 20mM glycine for 10 min at RT, followed by blocking with PBS containing 2 % BSA for 30 min at RT. Virus infected cells was stained for 1 h with anti-SARS-CoV-2 N protein rabbit monoclonal antibody (Sino Biological 40143-R001) diluted 1:1000 in blocking buffer, followed by secondary donkey anti-rabbit IgG (H+L) Alexa Fluor 488 antibody (Invitrogen) 1:1000 in blocking buffer for 30 min and DAPI staining (0.1 ug/mL in PBS) for 5 min. Number of infected cells were quantified using a TROPHOS Plate RUNNER HD^®^ (TROPHOS SA, Marseille, France).

### Statistics

Statistical analysis was performed by using Prism 8 (Graphpad software). Two-way ANOVA was used to determine the potential significance of detection of COVID-19 samples, as compared with controls. Correlative analysis was done by calculation of the Spearman r correlation.

## RESULTS

### Validation of an S-based assay for detection of early seroconversion during COVID-19 by venous blood draws

Our first aim was to develop a sensitive assay to detect anti-SARS-CoV-2 S IgG from venous blood. We therefore produced recombinant trimeric S proteins (S-2P) from the SARS-CoV-2 isolate Wuhan Wu-1(16) and used these as antigens to develop an ELISA-based set-up for detection of anti-S-directed IgG in serum or plasma of SARS-CoV-2 infected individuals (S-ELISA). After confirmation that we could detect anti-S IgG in a concentration-dependent manner (**Supplemental figure 1A**), we proceeded to define a cut-off for positive signal when used in single dilution-point format against 144 pre-pandemic samples and 30 samples from COVID-19+ individuals. The mean OD value of pre-pandemic samples was 0.06 +/-0.12 and we set a stringent cut-off for positive detection of S-specific IgG to OD 0.7 or higher (OD 0.06 + 5.8 StDev). By applying this cut-off, we detected seroconversion in 1/5, 6/12 and 13/13 PCR+ SARS-CoV2 infected individuals at 1-5, 6-14 or >14 days after disease onset, respectively **(Figure 1A)**. This accounted for 77% of all COVID-19 samples, irrespective of day of sampling. As expected, we observed a correlation between binding and neutralization **(Figure 1B)**. In addition, we showed that our assay had markedly increased sensitivity to detect low levels of S-specific IgG in serum samples, as compared with the (at the time) frequently used commercial assays Abbot Architect SARS-CoV-2 IgG or LIASON^®^ SARS-CoV-2 S1/S2 (**Supplemental figure 1B-D)**. As a final test of sensitivity, we screened 145 blinded serum samples from a Swedish care home for elderly with confirmed COVID-19 cases for the presence of anti-S IgG. Here, we observed seroconversion to anti-S IgG in 17 of 145 (11.7%) individuals, whereas the Abbot Architect SARS-CoV-2 assay only detected seroconversion to anti-N IgG in 10 (6.8%) individuals **(Figure 1C)**. Since binding to S-2P in serum from the 7 discrepant samples was abrogated by pre-incubation with an excess of soluble S-2P **(Figure 1D)**, we concluded that the 7 discrepant samples were from de-facto SARS-CoV-2 exposed individuals. These results were in line with a head-to-head comparison against 11 commercially available assays that were performed by the Swedish Public Health Agency, where our method (there, denominated *in house* RV (County of Västerbotten) had the highest sensitivity and specificity of all assays tested(17).

**Figure 1.**
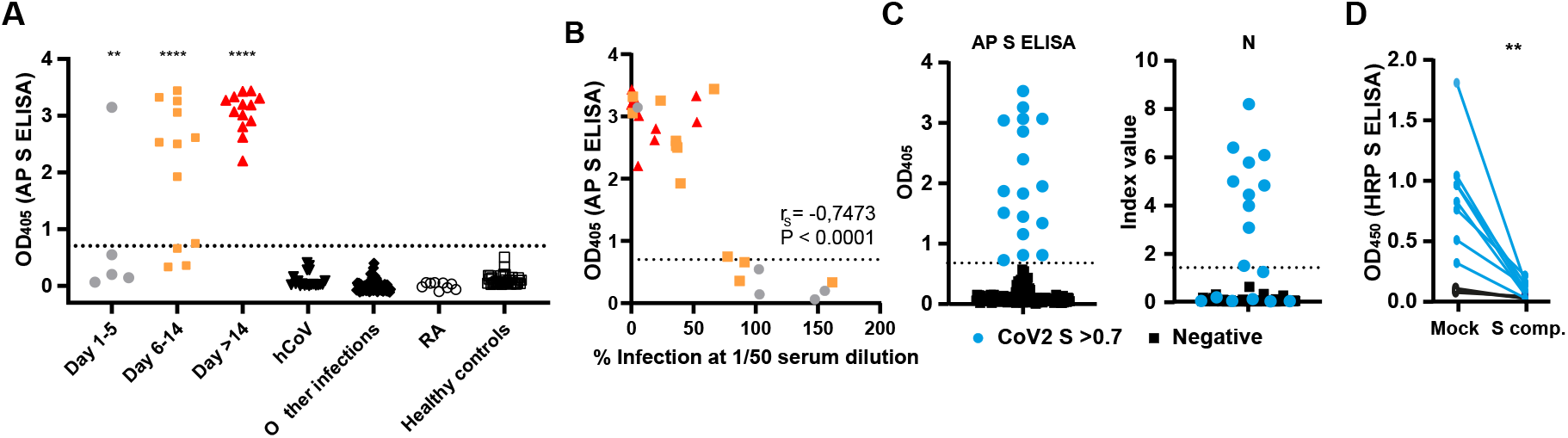
Detection of Anti-SARS-CoV-2 S-binding IgG and neutralizing antibodies. **A**. Single point validation of AP S ELISA set up. Shown are qPCR confirmed Covid-19 1-5 days post disease onset (n=5), 6-14 days (n=12), >14 days (n=13), hCoV (n=18), other viral infections (n=55), rheumatoid arthritis (RA) (n=9), and healthy controls (n=43). Dotted lines indicate threshold to define a positive sample (OD=0.7) **B**. Correlation between S ELISA and microneutralization at serum dilution 1/50. **C**. Comparison between S ELISA and Abbott Architect SARS-CoV-2 IgG (N) for detection of SARS-CoV-2 exposure. Blue circles indicate individuals positive for anti-S IgG in the S ELISA. Dotted line indicate threshold to define a positive sample (OD=0.7) **E**. Detection of anti-S IgG in serum that has been co-incubated with an excess of soluble S-2P protein.

### Capillary blood sampling followed by high quality detection of anti-S IgG by S-ELISA

After validating our assay on venous blood samples in a clinical setting, it was implemented as a benchmark assay at Region Västerbotten for serological assessment of previous COVID-19-exposure. Västerbotten is a large but sparsely populated region in northern Sweden. To facilitate equal access to high quality serological assessment for the population, we proceeded to clinically validate the use of the S-ELISA on capillary blood samples (quantitative dried blood spots, qDBS). We first performed parallel venous and qDBS blood sampling on 65 individuals with confirmed SARS-CoV-2 exposure and 84 individuals without previous exposure to the virus and then compared both sampling methods. The results verified that S-specific antibodies could be eluted from the dried blood spots, and that these could bind S-2P in a concentration-dependent manner (**Figure 2A**). In addition, we found a strong linear relationship between OD values between venous and qDBS sampling (r^2^=0.96; p<0.0001, **Figure 2B**), and that binding was specific to S-2P (Supplemental **figure 2A**). To investigate the possibility of using qDBS sample cards for at-home-sampling, we then tested if samples were affected by storage under summer of winter conditions prior to analysis. This was done by analysis of S-specific IgG from qDBS sample cards that had been incubated under artificial winter or summer conditions for 56h. The qDBS sample cards had been spiked with SARS-CoV-2 positive (N=8) or negative plasma (N=2). Results from this experiment demonstrated that all positive samples were well above background and that results were similar for all but one individual sample, where the qDBS had been incubated under summer conditions (**Figure 2C**). However, the majority of qDBS were unaffected and deemed suitable for at-home-sampling. As a final test, we performed a survey to understand the ease by which qDBS blood sampling could be managed by unexperienced individuals. In this survey, we found that only 3% of individuals reported difficulties with the sampling (**Supplemental figure 2B**). Collectively, these results validated an at-home-based strategy for S-based serology in Region Västerbotten, Sweden.

**Figure 2.**
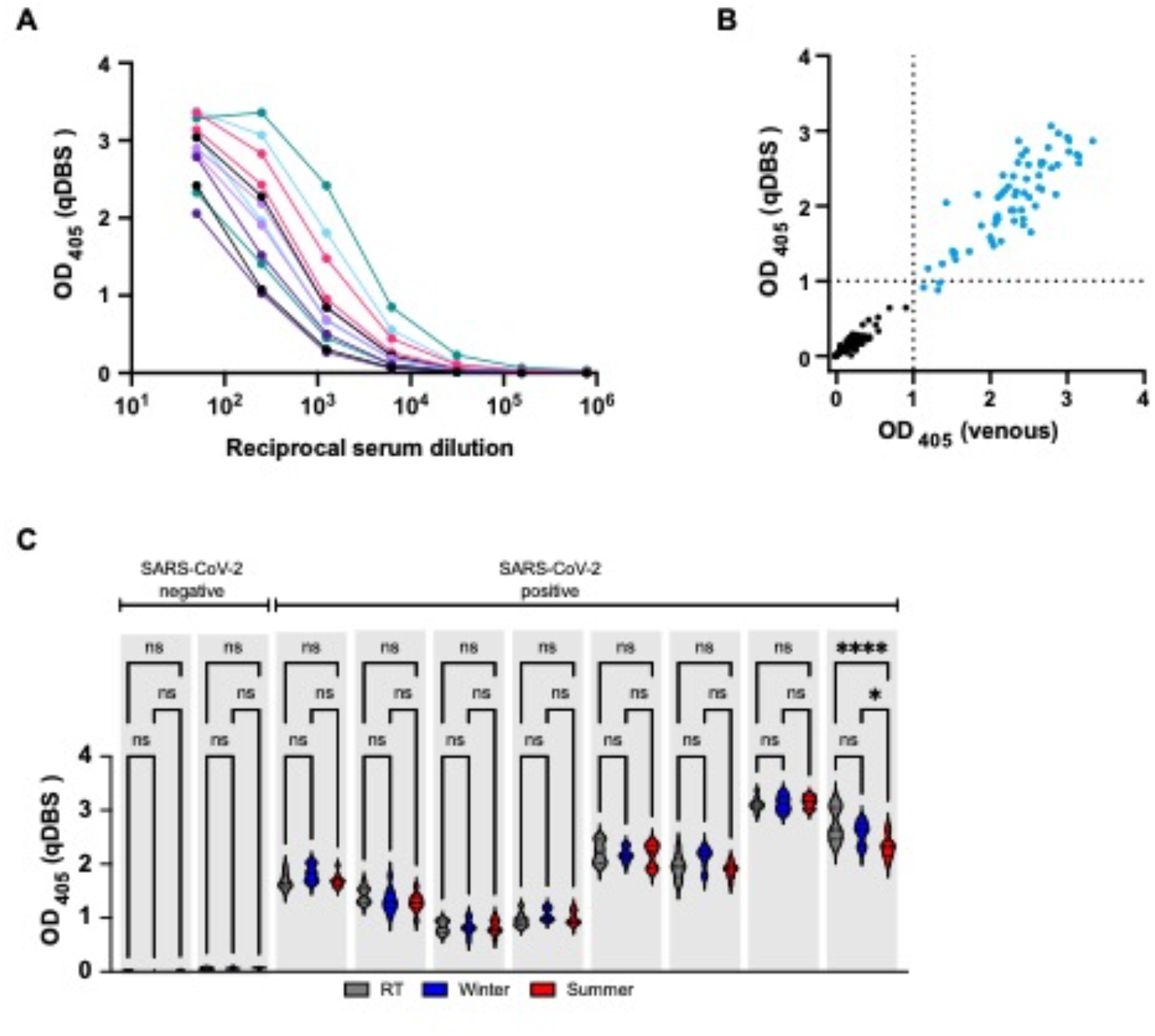
Verification of at-home testing methodology. **A**. The binding of antibodies extracted from eleven individuals sampled with quantitative dried blood spots (qDBS) sample cards to S-2P in concentrations 1/50 to 1/718250. **B**. Comparison of ELISA optical density readout of samples from 65 SARS-CoV2 experienced and 84 naïve individuals obtained from qDBS sample cards and venous sampling. **C**. Analysis of serum from ten individuals extracted from qDBS cards stored under conditions that mimic extreme summer or winter. *= p<0.05 ****= p<0.0001.

### Individually driven SARS-CoV-2 serology by self-sampling in a rural region of Sweden

Voluntary at-home-sampling in combination with analysis with S-ELISA at the hospital laboratory was implemented at Region Västerbotten during March to June 2021. In total, 4122 individual tests were distributed throughout the region and 91% were returned to the laboratory for analysis (**Figure 3A**). Of these, 96.7% were of adequate quality for elution of proteins from the dried blood spots (Figure 3B). The distribution of tests was relatively even between genders and most tests had been requested from individuals between 30-59 years of age (**Figure 3C**). During the test period, there had been an increase of individuals diagnosed with SARS-CoV-2 infection by PCR. Consistently, we could demonstrate an increase in frequency of S-positive qDBS samples that had been analyzed (Figure 3D).

**Figure 3.**
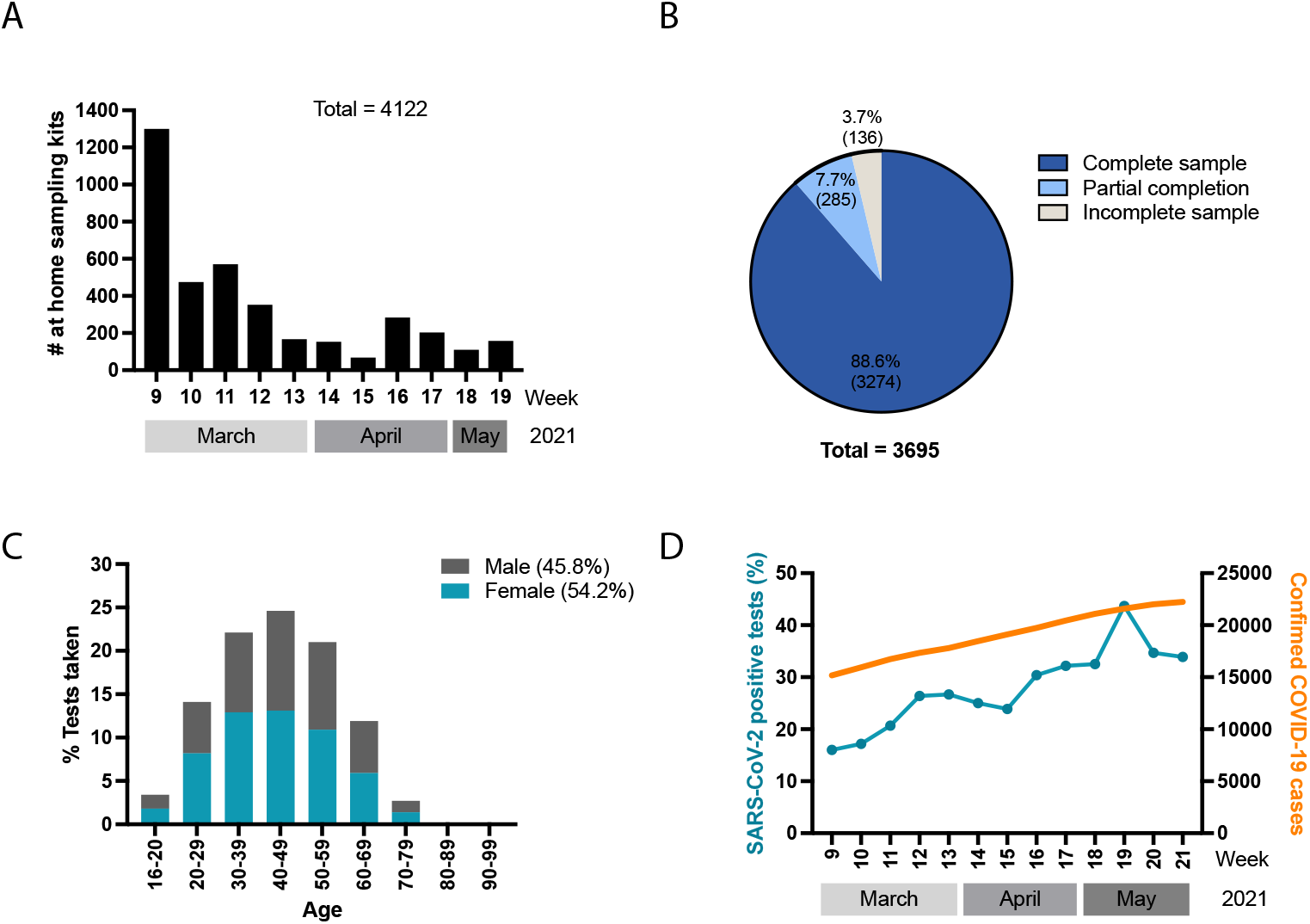
At-home testing of the population of Västerbotten. **A**. Shows the amount of quantitative dried blood spots (qDBS) tests distributed over time in Västerbotten during March to June 2021. **B**. Pie chart representing the success rate of qDBS tests returned for analysis. The dark blue field represents the amount of qDBS cards returned with two out of two chambers filled with blood. The light blue represents cards with one of two chambers filled with blood. The light gray area represents failed tests and the dark gray area represents tests that were not returned. **C**. Chart of age and gender distribution of successfully assayed tests. **D**. The fraction of serological tests found to be positive for anti-S IgG (blue dotted line) and the accumulated number of SARS-CoV-2-positive qPCR tests (orange line) in Västerbotten during the same period of time.

Before the implementation of at-home-sampling, inhabitants of the County of Västerbotten region had been offered serological testing using venous sampling followed by analysis of N- and S-specific IgG. This sampling had been performed by healthcare professionals during November 2020 through January 2021. A total of 8002 venous blood draws had been performed at health care centers or mobile sampling centers. This allowed us to compare the geographical distribution of venous blood sampling with at-home-sampling. By this, we found that the at-home-sampling strategy had resulted in an increased proportion of tests in rural areas while the strategy of venous blood sampling was skewed towards the two major urban areas of Umeå and Skellefteå. (**Figure 4**).

**Figure 4.**
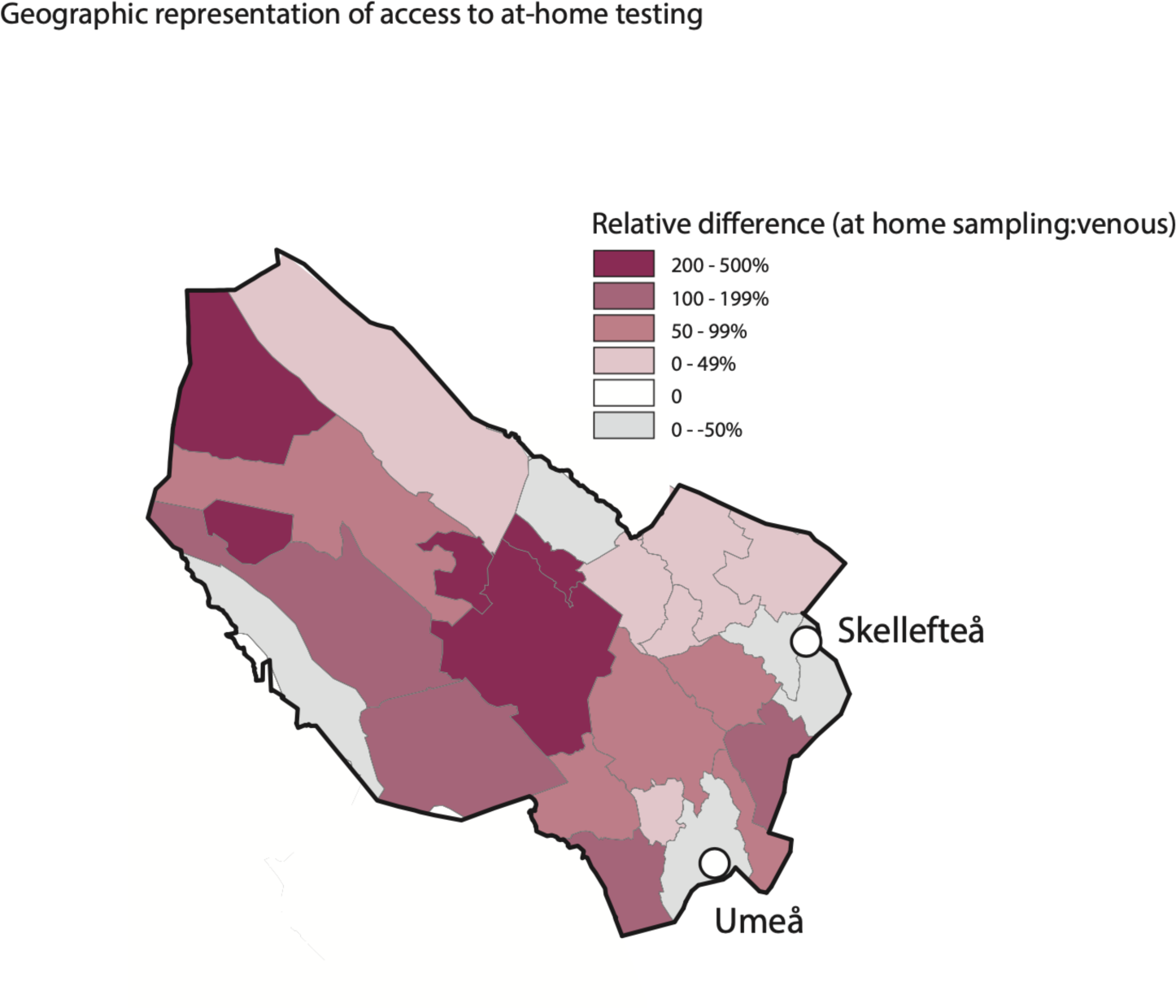
Access to serological testing in Västerbotten. A map representation of Västerbotten subdivided in geographical area codes. The color distribution shows the over or under representation of at-home testing using quantitative dried blood spots (qDBS) in relationship to venous testing for serology. The two urban areas Umeå and Skellefteå are indicated with circles.

## Discussion

In this study, we outline the development and clinical implementation of a sensitive method for detection of SARS-CoV-2 exposure and the further development of this assay for at-home sampling. The rationale for the latter was to increase access to high quality diagnostics for all individuals within a large but sparsely populated region of Sweden.

As demonstrated by us, and by independent studies by the Swedish Public Health Agency, our assay for detection of S-specific IgG had the highest standard with regards to sensitivity and specificity (17). For this reason, the assays was subsequently used on venous blood samples for probabilistic classification of anti-SARS-CoV-2 antibody responses (18).

When applying eluates from qDBS sample cards instead of serum from venous blood draws, we could demonstrate a high correlation between the sampling methods. Importantly, the detection of S-specific IgG from qDBS elutes were minimally affected by conditions that mimic climate variations in the northern of Sweden. Here, the strategy was implemented in clinical care and over 4000 tests were distributed throughout the region and then analyzed at the clinical laboratory at Norrland’s University Hospital.

The strategy outlined herein has several benefits to that of serological analysis by venous blood sampling. These include: sampling that do not require trained professionals, outreach to geographically challenging areas and reduced risk for spread of SARS-CoV-2 upon sampling. Moreover, the use of qDBS allow for downstream processing and implementation of tests of with high specificity and sensitivity(19-21),.

Collectively, we here present a strategy for serological surveillance that was implemented for serosurveillance of SARS-CoV-2 infection by at-home sampling. We show that this method can be implemented in clinical care even during winter season in arctic or subartic areas. The approach increased the possibility of individuals in rural regions to test for previous SARS-CoV-2 exposure. This methodology is also possible to directly implement as a practical tool to monitor vaccine-induced responses to the S-protein of SARS-CoV-2 in large populations.

## Data Availability

Data available within the article or in supplementary materials

## Acknowledgements

We would like to acknowledge the Department of Clinical Microbiology, the Department of Infection Prevention and Control and the Department of Infectious Diseases, Region Västerbotten, Sweden and Umeå Center for Microbial Pathogenesis, and the Translational Research Center Umeå, Sweden for facilitating the research. We also like to acknowledge Bert Blomqvist and Ingrid Marklund at the clinical microbiology laboratory at Region Västerbotten.

Funding was through grants from Vinnova (2020-03103) to Xerum AB and MF, The Swedish Research Council (2020-06235), Umeå University (FS 2.1.6-1233-20) and from the SciLifeLab National COVID-19 Research Program (C19VE:007), financed by the Knut and Alice Wallenberg Foundation to MF; the Swedish Research Council (2021-04665) and Region Västerbotten (REF) to CA. J.N. is a Wallenberg Center for Molecular Medicine Associated Researcher.

## Conflict of Interest

JWB, JN and MF are founders and share-holders of the diagnostic company Xerum AB.

---

**Supplemental figure 1.**
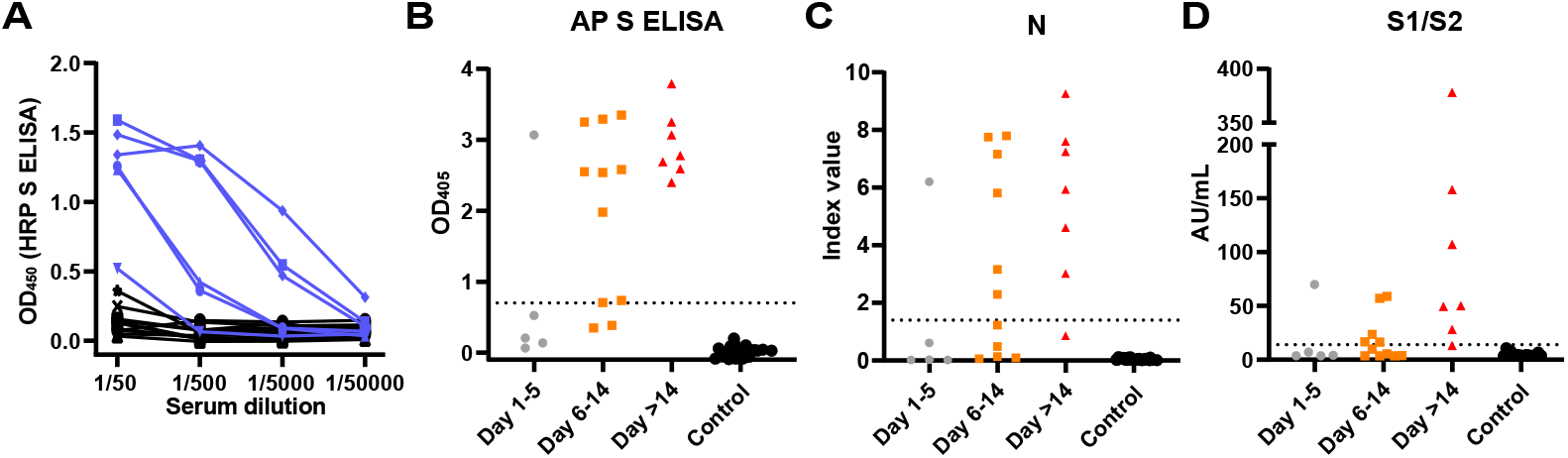
**(A)** Anti-S IgG in serum from COVID-19 patients or none-exposed individuals at different serum dilutions. Comparison for detection of previous SARS-CoV-2 exposure at different timepoints after disease onset in COVID-19 patients or healthy controls. Shown is **(B)** S ELISA (S-2P based), **(C)** Abbott Architect SARS-CoV-2 IgG (N-based) or **(D)** LIASON^®^ SARS-CoV-2 S1/S2 (wt S-based).

**Supplemental figure 2.**
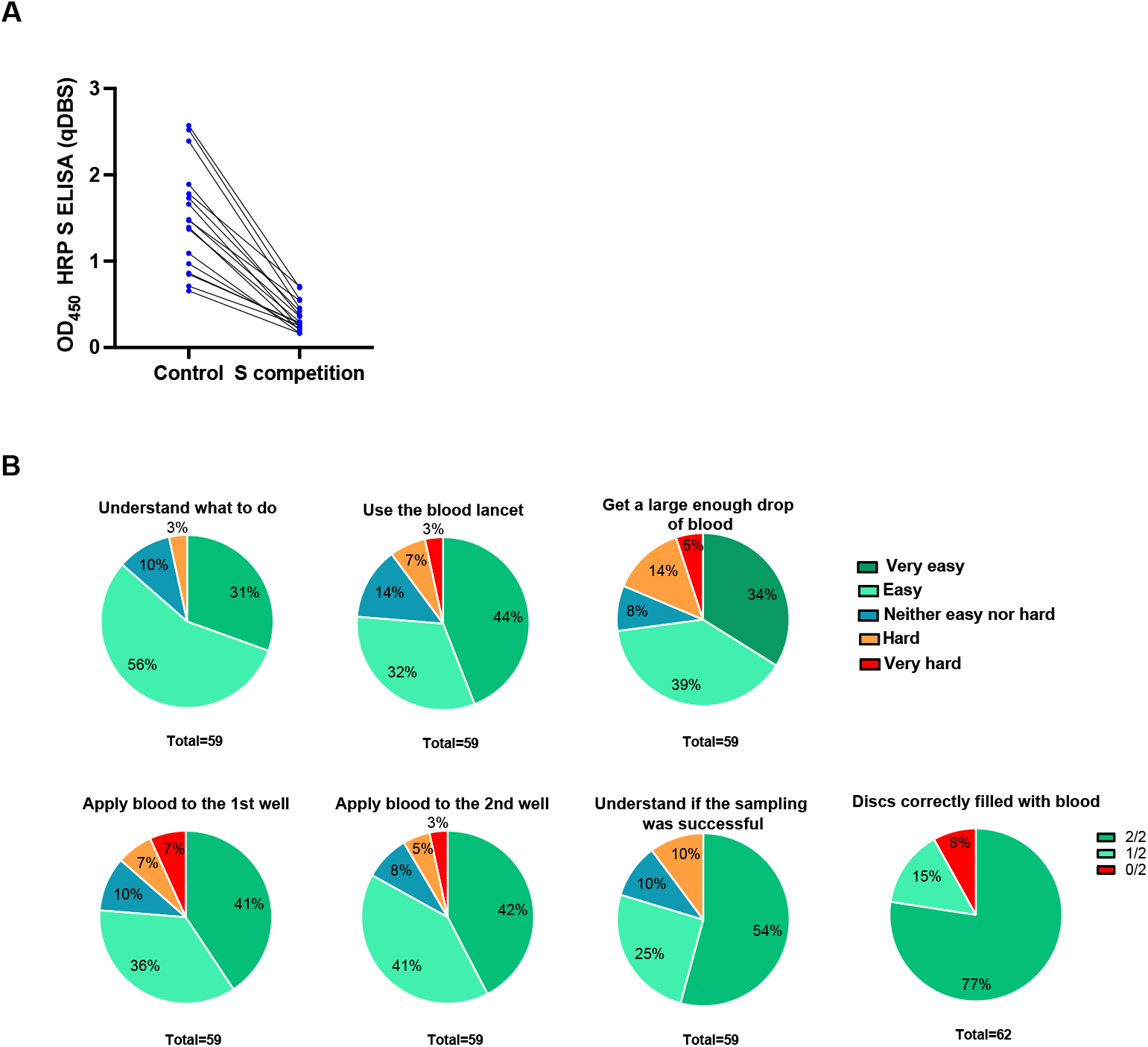
**(A)** Detection of anti-S IgG in qDBS elutes after co-incubation with an excess of soluble S-2P protein. (B) Evaluation of the ease of self-sampling with the provided self-sampling kit.

## Notes

### Competing Interest Statement

JWB, JN and MF are founders and share-holders of Xerum AB.

### Funding Statement

Funding was through grants from Vinnova (2020-03103) to Xerum AB and MF, The Swedish Research Council (2020-06235), Umea University (FS 2.1.6-1233-20) and from the SciLifeLab National COVID-19 Research Program (C19VE:007), financed by the Knut and Alice Wallenberg Foundation to MF; the Swedish Research Council (2021-04665) and Region Vasterbotten (REF) to CA. J.N. is a Wallenberg Center for Molecular Medicine Associated Researcher.

### Author Declarations

Ethical permission for the study was obtained from the Swedish Ethical review authority (No: 2020-01557) and all research was carried out according to The Code of Ethics of the World Medical Association (Declaration of Helsinki).

### Summary of Updates

We here update the manuscript and title according to further experiments to adapt our methodlogy to at-home sampling.

## References

1. Milne G, Hames T, Scotton C, Gent N, Johnsen A, Anderson RM, et al. Does infection with or vaccination against SARS-CoV-2 lead to lasting immunity? Lancet Respir Med. 2021;9(12):1450–66.

2. Altarawneh HN, Chemaitelly H, Hasan MR, Ayoub HH, Qassim S, AlMukdad S, et al. Protection against the Omicron Variant from Previous SARS-CoV-2 Infection. N Engl J Med. 2022;386(13):1288–90.

3. Cox RJ, Brokstad KA. Not just antibodies: B cells and T cells mediate immunity to COVID-19. Nat Rev Immunol. 2020;20(10):581–2.

4. Krammer F, Simon V. Serology assays to manage COVID-19. Science. 2020;368(6495):1060–1.

5. Lopman BA, Shioda K, Nguyen Q, Beckett SJ, Siegler AJ, Sullivan PS, et al. A framework for monitoring population immunity to SARS-CoV-2. Ann Epidemiol. 2021;63:75–8.

6. Shioda K, Lau MSY, Kraay ANM, Nelson KN, Siegler AJ, Sullivan PS, et al. Estimating the Cumulative Incidence of SARS-CoV-2 Infection and the Infection Fatality Ratio in Light of Waning Antibodies. Epidemiology. 2021;32(4):518–24.

7. To KK, Tsang OT, Leung WS, Tam AR, Wu TC, Lung DC, et al. Temporal profiles of viral load in posterior oropharyngeal saliva samples and serum antibody responses during infection by SARS-CoV-2: an observational cohort study. Lancet Infect Dis. 2020;20(5):565–74.

8. Long QX, Liu BZ, Deng HJ, Wu GC, Deng K, Chen YK, et al. Antibody responses to SARS-CoV-2 in patients with COVID-19. Nat Med. 2020.

9. GeurtsvanKessel CH, Okba NMA, Igloi Z, Bogers S, Embregts CWE, Laksono BM, et al. An evaluation of COVID-19 serological assays informs future diagnostics and exposure assessment. Nat Commun. 2020;11(1):3436.

10. Wang Y, Wang Y, Chen Y, Qin Q. Unique epidemiological and clinical features of the emerging 2019 novel coronavirus pneumonia (COVID-19) implicate special control measures. J Med Virol. 2020.

11. Ing AJ, Cocks C, Green JP. COVID-19: in the footsteps of Ernest Shackleton. Thorax. 2020:thoraxjnl-2020-215091.

12. Okba NMA, Muller MA, Li W, Wang C, GeurtsvanKessel CH, Corman VM, et al. Severe Acute Respiratory Syndrome Coronavirus 2-Specific Antibody Responses in Coronavirus Disease 2019 Patients. Emerg Infect Dis. 2020;26(7).

13. Hoffmann M, Kleine-Weber H, Schroeder S, Kruger N, Herrler T, Erichsen S, et al. SARS-CoV-2 Cell Entry Depends on ACE2 and TMPRSS2 and Is Blocked by a Clinically Proven Protease Inhibitor. Cell. 2020;181(2):271–80 e8.

14. Jiang S, Hillyer C, Du L. Neutralizing Antibodies against SARS-CoV-2 and Other Human Coronaviruses. Trends Immunol. 2020;41(5):355–9.

15. Wrapp D, Wang N, Corbett KS, Goldsmith JA, Hsieh CL, Abiona O, et al. Cryo-EM structure of the 2019-nCoV spike in the prefusion conformation. Science. 2020;367(6483):1260–3.

16. Amanat F, Stadlbauer D, Strohmeier S, Nguyen THO, Chromikova V, McMahon M, et al. A serological assay to detect SARS-CoV-2 seroconversion in humans. Nat Med. 2020.

17. Lagerqvist N, Maleki KT, Verner-Carlsson J, Olausson M, Dillner J, Wigren Bystrom J, et al. Evaluation of 11 SARS-CoV-2 antibody tests by using samples from patients with defined IgG antibody titers. Sci Rep. 2021;11(1):7614.

18. Castro Dopico X, Muschiol S, Grinberg NF, Aleman S, Sheward DJ, Hanke L, et al. Probabilistic classification of anti-SARS-CoV-2 antibody responses improves seroprevalence estimates. Clin Transl Immunology. 2022;11(3):e1379.

19. Klumpp-Thomas C, Kalish H, Drew M, Hunsberger S, Snead K, Fay MP, et al. Standardization of ELISA protocols for serosurveys of the SARS-CoV-2 pandemic using clinical and at-home blood sampling. Nat Commun. 2021;12(1):113.

20. Toh ZQ, Higgins RA, Anderson J, Mazarakis N, Do LAH, Rautenbacher K, et al. The use of dried blood spots for the serological evaluation of SARS-CoV-2 antibodies. J Public Health (Oxf). 2021.

21. Roxhed N, Bendes A, Dale M, Mattsson C, Hanke L, Dodig-Crnkovic T, et al. Multianalyte serology in home-sampled blood enables an unbiased assessment of the immune response against SARS-CoV-2. Nat Commun. 2021;12(1):3695.

